# Utilization of Youth-Friendly Reproductive Health Services and Associated Factors among Secondary School Students in Addis Ababa, Ethiopia

**DOI:** 10.1101/2025.02.25.25322847

**Authors:** Yitbarek Biru Huluka, Tamirat Tesfaye Dasa

**Affiliations:** Rift Valley University, School of Postgraduate Studies, P.O. Box 80734, Addis Ababa, Ethiopia; Hawassa University, College of Medicine and Health Science, P.O. Box 1560, Hawassa, Ethiopia

**Keywords:** Reproductive Health, youths, secondary school, Yeka Sub-City, Ethiopia

## Abstract

**Introduction:** Due to disparities in data gathering and dissemination between nations, it is challenging to provide exact statistics. However, the global scope of reproductive health problems within schools is substantial, with numerous problems influencing the development and well-being of young people.

**Objective:** This study aimed to assess the utilization of youth-friendly reproductive health services (YFRHS) by secondary school students in Yeka Sub-City, Addis Ababa, Ethiopia.

**Method:** To examine the YFRHS among secondary school students in Yeka Sub-City, Addis Ababa, an institution-based cross-sectional survey was conducted between June and July 2024. A total of 415 students from 15 secondary schools who were chosen randomly participated in this study. A standard, well-structured, self-administered questionnaire was used to collect the data. After cleaning, coding, and importing into EpiData version 4.6.0.2, data were exported to SPSS Windows version 27. Factors related to YFRHS use were identified using binary logistic regression analysis. In multivariable logistic regression, every factor in the bivariate analyses, with a p-value less than 0.25 in the bivariate analyses was included. The strength of relationship was evaluated using the adjusted odds ratio and a 95% CI, with a P value ≤ 0.05 was taken into consideration to announce the statistical significance.

**Results:** Youth-friendly reproductive health services (YFRHS) utilization in this study was 18.8% (95% CI: 0.154, 0.227). This study found that having a sexual partner Adjusted Odd Ratio (AOR) (AOR: 3.8 (1.7, 8.4)), having a conversation with parents (AOR: 2.2 (1.2, 4.02)), having YFRHS facilities nearby (AOR: 5.1 (2.5, 10.3)), and believing that seeking behavior improves youth health (AOR: 4.6 (1.5, 14.04)) were all significantly associated with reproductive health utilization.

**Conclusion:** Among school-aged youth, one in five used youth-friendly reproductive health services. Reproductive health use was significantly associated with having a sexual partner, having a favorable perspective on whether seeking behavior benefits youth health, having discussions with parents, and having YFRHS facilities in the living area.

## Background

Youths are people aged between 15 and 24 years old. Globally, there were 37.7 million people live with HIV/AIDS, and 90% occur among youths. In today’s rapidly evolving world, youth well-being and empowerment are essential for building a healthy and prosperous society. Central to this is the provision of comprehensive reproductive health services that cater to adolescents’ unique needs of adolescents (1).

Adolescents are at a critical stage in their lives when they experience physical, emotional, and social changes, which make them particularly vulnerable to reproductive health challenges (2). In many cases, accessing accurate information and appropriate healthcare services can be challenging owing to societal taboos, a lack of awareness, or limited resources. Integrating reproductive health services within schools provides a safe and supportive environment for young people to seek the guidance and care they need (3).

Schools play a unique and influential role in shaping the lives of students. By introducing comprehensive reproductive health services, educational institutions can ensure that young people receive accurate information, guidance, and support from qualified professionals (4).

Students are the most vulnerable group to reproductive health problems because of their inclination to engage in risky sexual behaviors. Pregnancy among adolescents remains a major concern worldwide. According to the World Health Organization (WHO), an estimated 21 million girls aged 15 _ 19 years become pregnant in developing regions annually. Pregnancies often have adverse health, social, and economic consequences for young mothers and their children (5).

Reproductive health issues in schools have profound mental health implications for young people. Stress, anxiety, and social stigma associated with unintended pregnancies, STIs, and challenges related to sexual orientation and gender identity contribute to mental health disorders and emotional distress (6).

Young people aged 10 to 24 comprise the largest number of Ethiopians, accounting for 33.8% of the population. However, many youths are less informed about reproductive health services and are less comfortable accessing these services than are adults (7). The effects of these efforts are not well understood across Ethiopian high schools and preparatory schools as evidenced by persistent reproductive health problems and challenges to the youths (8).

Advocacy, awareness and educational programs to support the utilization of RH services are beginning to spread. Some clinical records indicate that most young people increasingly attend RH clinics to access contraceptives and condoms. However, statistics show that the rates of STIs and unwonted pregnancies among adolescents remain high in Addis Ababa (9). Adolescents and youth still face RH challenges as unsafe abortion, unmet need for family planning and STI including HIV. Furthermore, the knowledge and utilization of youth-friendly reproductive health services (YFRHS) in available locations have not been assessed in an all-inclusive manner. This study aimed to determine adolescents’ knowledge of reproductive health and, utilization of youth-friendly services: their knowledge of the components of the services.

## Methods and Materials

### Study design and period

An institution-based cross-sectional study was conducted to assess reproductive health utilization of the YFRHS among secondary school students in Yeka Sub-City, Addis Ababa. The study was conducted from June to July 2024.

### Study area

This study was conducted in Addis Ababa. Addis Ababa is the political, capital and the most important commercial and cultural center of Ethiopia, and is geographically located at the heart of the nation at, 9^0^ 2_’N latitude and 38^0^ 45’ E longitudes. Its average altitude is 2,400 m above sea level, with the highest elevation at Entoto Hill to the north, reaching 3,200 m with an annual growth rate of 2.1%. The population of Adolescents comprises 29.1% of the total population. The city is divided into 11 administrative sub-cities and has 116 health centers, of which 86 are governmental and the rest are owned by non-governmental organizations (NGOs). Addis Ababa has, 52 Hospitals (13-governmental, 35-private and 4-NGOs) and 534 clinics, of which 34 are owned by NGOs (10). Yeka Sub-City is geographically located in the northeastern part of Addis Ababa, which has a dynamic topography. It borders the Gullele, Arada, Kirkos and Bole Sub-Cities. It covers 85.98 KM^2^ Area with population projection (2022) of 488.537; of which 225,543 (46.2%) were males and the remaining 262,994 (53.8%) were females. It has 15 secondary schools, including both public and private sectors, with 17,704 secondary school students.

### Target population

All secondary school youths (15–24) years old were studying in secondary schools in Yeka Sub-City, Addis Ababa was the target population in this study.

### Study population

Randomly selected youth students aged 15-24 years old were enrolled in the secondary schools of Yeka Sub-City, Addis Ababa during the study period

### Inclusion and exclusion criteria

#### Inclusion criteria

Youths in the age group of 15-24 years old studying in secondary schools of Yeka Sub-City and who provided written informed assent for participants aged 15-18 years old were secured from their guardians/parents and consent for participants aged 19-24 years old were included in this study.

#### Exclusion criteria

Youths (15-24 years old) who were unable to participate in the study due to mental or severe physical illnesses were excluded from the study.

### Sample size determination

A single_ population proportion formula was used to estimate the sample size. From the study conducted in Addis Ababa taking health service utilization as p=42.9 % study done in the previous study, Addis Ababa (11) was used to calculate the sample size.

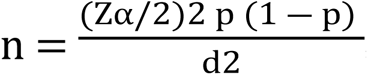, Where: Zα/2 is = the given confidence level value, which is 1.96

P= 42.9% proportion of service utilization

Maximum acceptable difference (d=0.05) =3.8416*0.429(0.571)/0.05*.05

n= (1.962(0.429) (1–0429))/ ((0.05)2) =376.41

Adding 10% of (376.41) non-response rate =37.64

n =376.41+37.64 = 414.05 =415

Therefore, the maximum sample sizes of 415 participants were included in the study.

### Sampling Technique and Procedure

**Figure 1:**
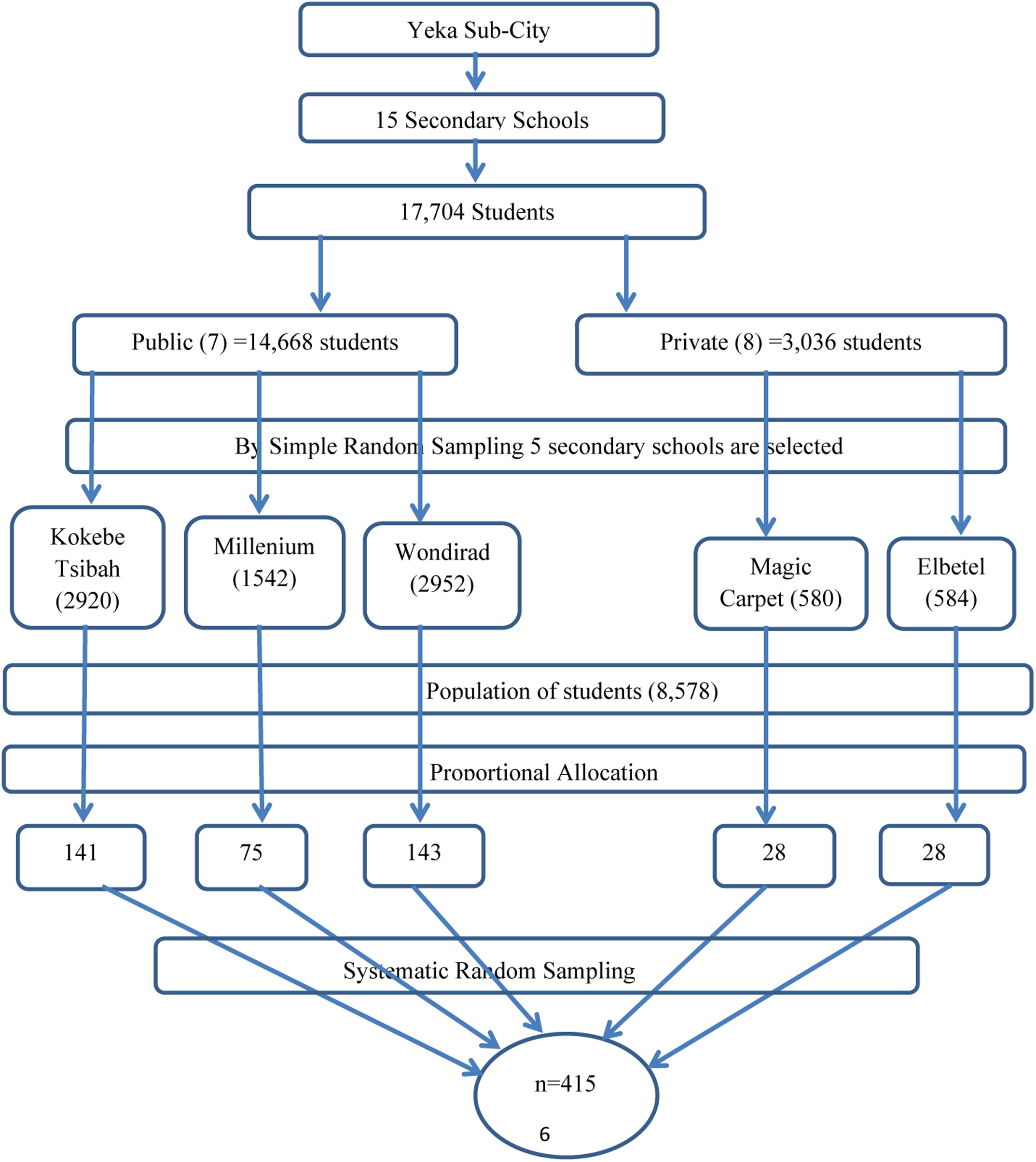
Steps of sampling procedure and sampling technique.

#### Variables

##### Dependent variable

Utilization of youth-friendly reproductive health services (Yes/No)

### Independent variables

The independent variables in this study were demographic factors such as age, sex, educational status, socio-economic factors and school factors, such as the; educational and employment status of the parents/household heads. Socio-cultural factors included religion. Health system factors included health facility organization and service delivery, health provider attitude, and availability of youth-friendly health services within the study area.

### Operational Definitions

#### Youth-Friendly Reproductive Health Service

Incorporates, general counseling services, family planning service, voluntary counseling and testing (VCT) for HIV, Condom use, treatment of sexually transmitted infections, perinatal care (Antenatal Care (ANC), delivery and postnatal care), abortion and post-abortion care services.

#### YFRHS Utilization

Utilization of at least one component of youth-friendly reproductive health services.

#### Youth Center

is a social and recreational center intended primarily for use by young people

### Data Collection Methods and Tools

#### Data collection instruments

Data were collected using a structured self-administered questionnaire developed in the literature (12). The questionnaire was translated into the local language (Amharic) and back_translated into English to ensure consistency. A Pre-test was conducted on 5% of the sample size out of the study area at Bole Beshale Secondary School, Lemi-Kura Sub-City. A necessary correction was made based on the results of the pre-test data analysis before the actual data collection.

### Data collection procedures

First, training was given the data collectors were trained regarding the study objectives, method of data collection tools for data collection, and procedures to be followed for data collection. Senior public health professionals were recruited to supervise data collection.

#### Data quality control

Individuals tasked with collecting and overseeing data were provided with comprehensive training for one day on the objectives of the study, effective interviewing techniques, proper questionnaire administration, and identification of skipping patterns. Prior to actual data collection, a pre-test was conducted on a 5% sample size in Bole Beshale, Lemi-Kura Sub-City, Addis Ababa, Ethiopia to assess the questionnaire’s clarity and sequence of questions. Based on the results, questions that were deemed unclear or confusing were refined. To ensure completeness, the supervisor checked the data daily during the data-collection period. Additionally, each questionnaire was assigned a unique identification number as a variable.

### Data Processing and Analysis

The collected data were cleaned, coded, entered into EpiData version 4.6.0.2, and exported to SPSS Windows version 27. Descriptive statistics were used to describe the percentage and number distributions of variables in the study. Continuous variables presented as a means and standard deviation (SD). Categorical variables presented as percentages. Binary logistic regression analysis was used to determine the factors associated with RH utilization separately. Bivariate logistic regression analysis was performed to examine the association of each factor with RH utilization. All factors p value < 0.25 in the bivariate analysis, were included in the multivariable logistic regression. Adjusted odds ratio (AOR) along with a 95% confidence interval was estimated to assess the strength of association, and a P value ≤ 0.05 were considered to indicate the statistical significance of in the multivariate analysis.

### Ethical Consideration

Ethical clearance was obtained from the Addis Ababa Public Health and Emergency Management Directorate and written permission was obtained from the corresponding district to collect data from the respective secondary schools. Participation in the study was voluntary, and participants were informed of their right not to participate in the study if they did not want to participate, and their right to withdraw from the study at any point during the interview. Written informed assent for participants aged 15-18 years old secured from their parents/guardians and consent for participants aged 19-24 years old was obtained from each participant after they were informed of the study’s purpose. Moreover, the confidentiality of the information was assured using an anonymous questionnaire. The respondents’ names and personal information were not included in the questionnaire. All documents were kept private and locked to maintain their confidentiality. Data were stored in a password-locked laptop.

## Results

### Socio-demographic Characteristics

A total of 415 respondents participated in the study, with a 100 % response rate. Two hundred twenty three (53.7%) female and 192 (46.3%) male students were participated. Approximately 359 (86.5%) public and 56 (13.5%) private school students had participated in this study. The majority nearly 390 (94 %) of the respondents were found below the age of 20, while the rest nearly 25(6%) were twenty and above with mean age and SD of 15, 23, and 17.11 with (±1.425 SD) respectively.

The majority 391 (94.2%) of the participants were single and 21 (5.1%) were married in marital status. In regard to the residence of parents 371 (89.4%) and 44 (10.6%) were urban and rural respectively. 114 (27.5%) grade nine, 113 (27.2%) grade ten, 88 (21.2 %%) Grade eleven and 100 (24.1%) Grade 12 students have participated in the study. The majority 351 (84.6 %) of the participant were living with their parents while 38 (9.2%) of them were living with their relatives. Approximately 163 (39.3%) and 252 (60.7%) of the participants had pocket money and had no pocket money, respectively.

The majority 391 (94.2%), of the study participants, were single and only 21 (5.1%) of them were married in marital status. The majority 360 (86.7%) of the participants were Orthodox in religion.

**Table 1:**
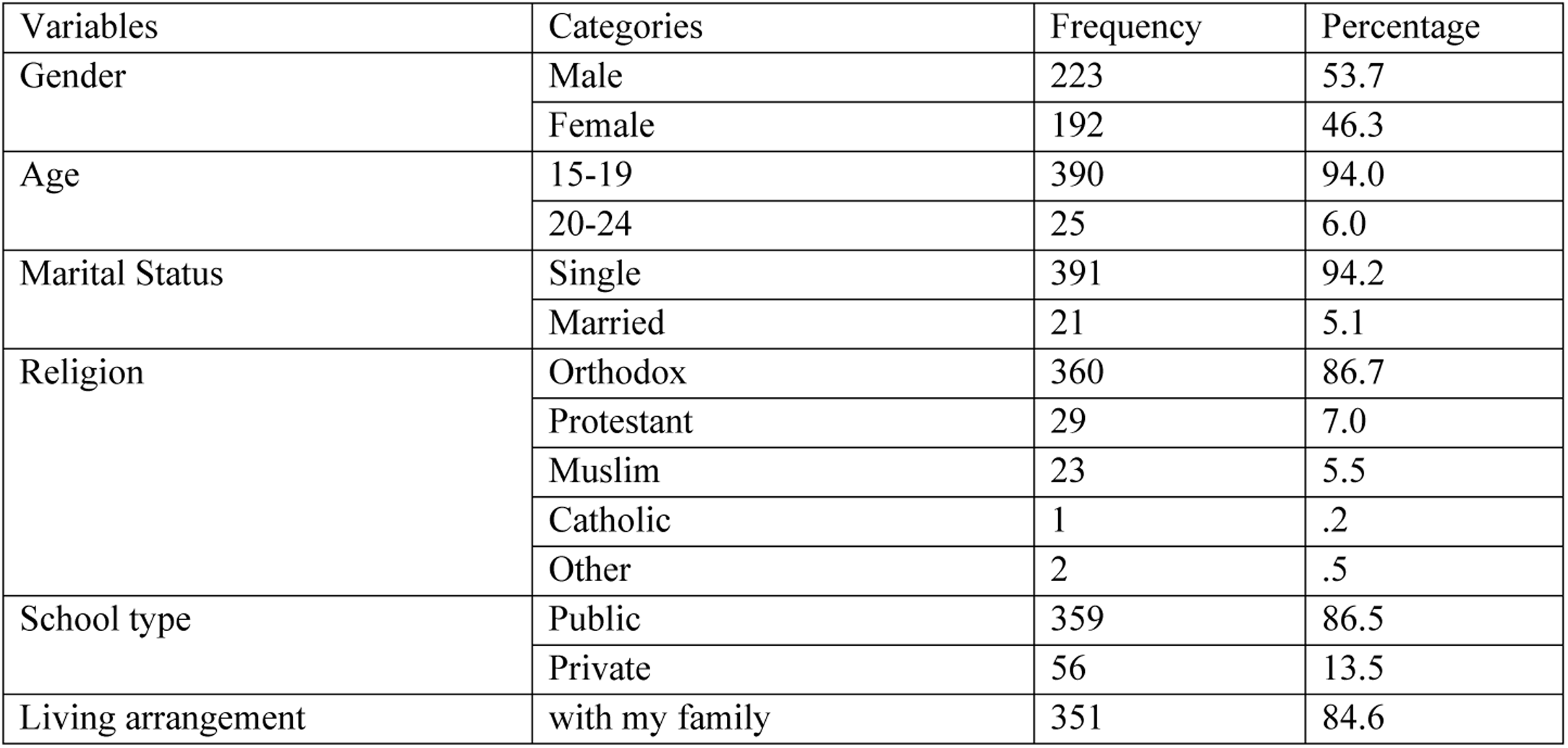
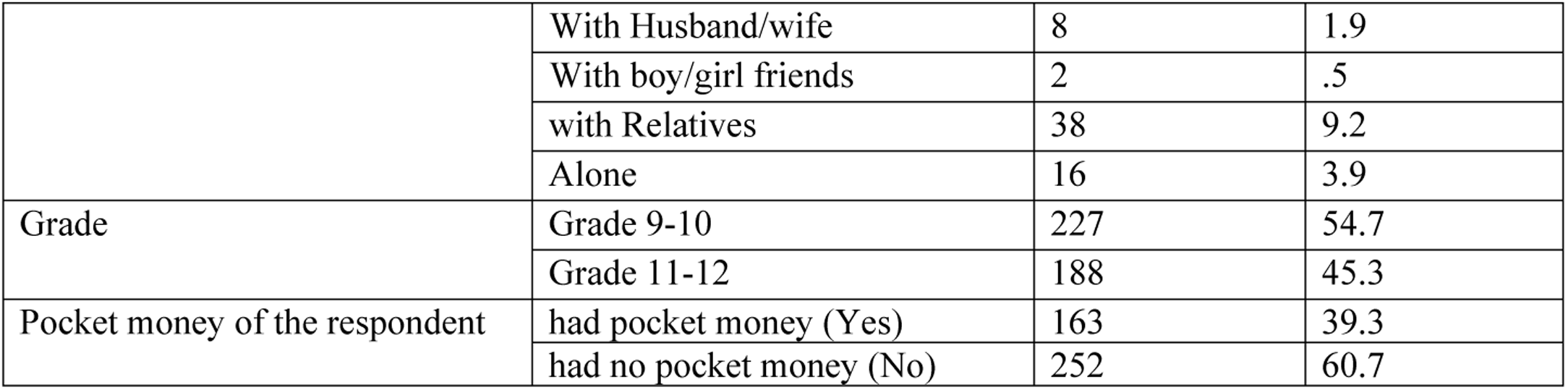
Respondents-related factors of YFRHS utilization, Yeka Sub-City, Addis Ababa, Ethiopia, 2024 (n=415)

#### Parent-related Socio-demographic characteristics

From the table shown below, the majority 371 (89.4%) of the parents’ residences were urban. And about two third 267 (64.3%) of fathers’ educational status were secondary and above level of education. Meanwhile, 219 (52.8%) of the participants’ mothers were secondary or above the level of education.

**Table 2:**
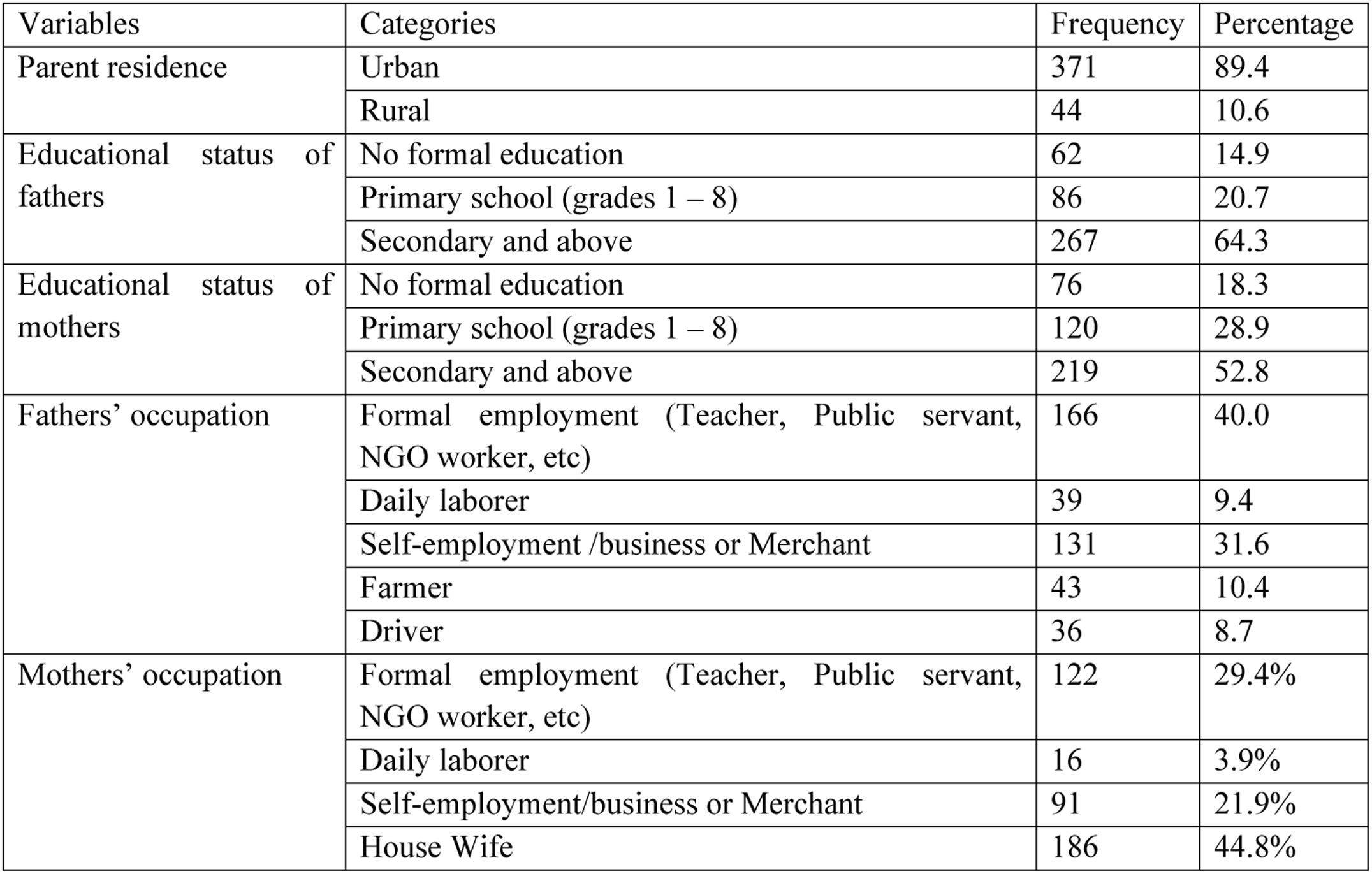
Parent-related factors of YFRHS utilization, Yeka Sub-City, Addis Ababa, Ethiopia, 2024 (n=415)

| Variables | Categories | Frequency | Percentage |
| --- | --- | --- | --- |
| Parent residence | Urban | 371 | 89.4 |
|  | Rural | 44 | 10.6 |
| Educational status of fathers | No formal education | 62 | 14.9 |
|  | Primary school (grades 1 – 8) | 86 | 20.7 |
|  | Secondary and above | 267 | 64.3 |
| Educational status of mothers | No formal education | 76 | 18.3 |
|  | Primary school (grades 1 – 8) | 120 | 28.9 |
|  | Secondary and above | 219 | 52.8 |
| Fathers' occupation | Formal employment (Teacher, Public servant, NGO worker, etc) | 166 | 40.0 |
|  | Daily laborer | 39 | 9.4 |
|  | Self-employment /business or Merchant | 131 | 31.6 |
|  | Farmer | 43 | 10.4 |
|  | Driver | 36 | 8.7 |
| Mothers' occupation | Formal employment (Teacher, Public servant, NGO worker, etc) | 122 | 29.4% |
|  | Daily laborer | 16 | 3.9% |
|  | Self-employment/business or Merchant | 91 | 21.9% |
|  | House Wife | 186 | 44.8% |

### Knowledge-related Variable of the Respondents

Two-thirds of 277 (66.7%) of the respondents had general information about youth reproductive health services while the rest 138 (33.3%) had no information about youth reproductive health services.

**Table 3:**
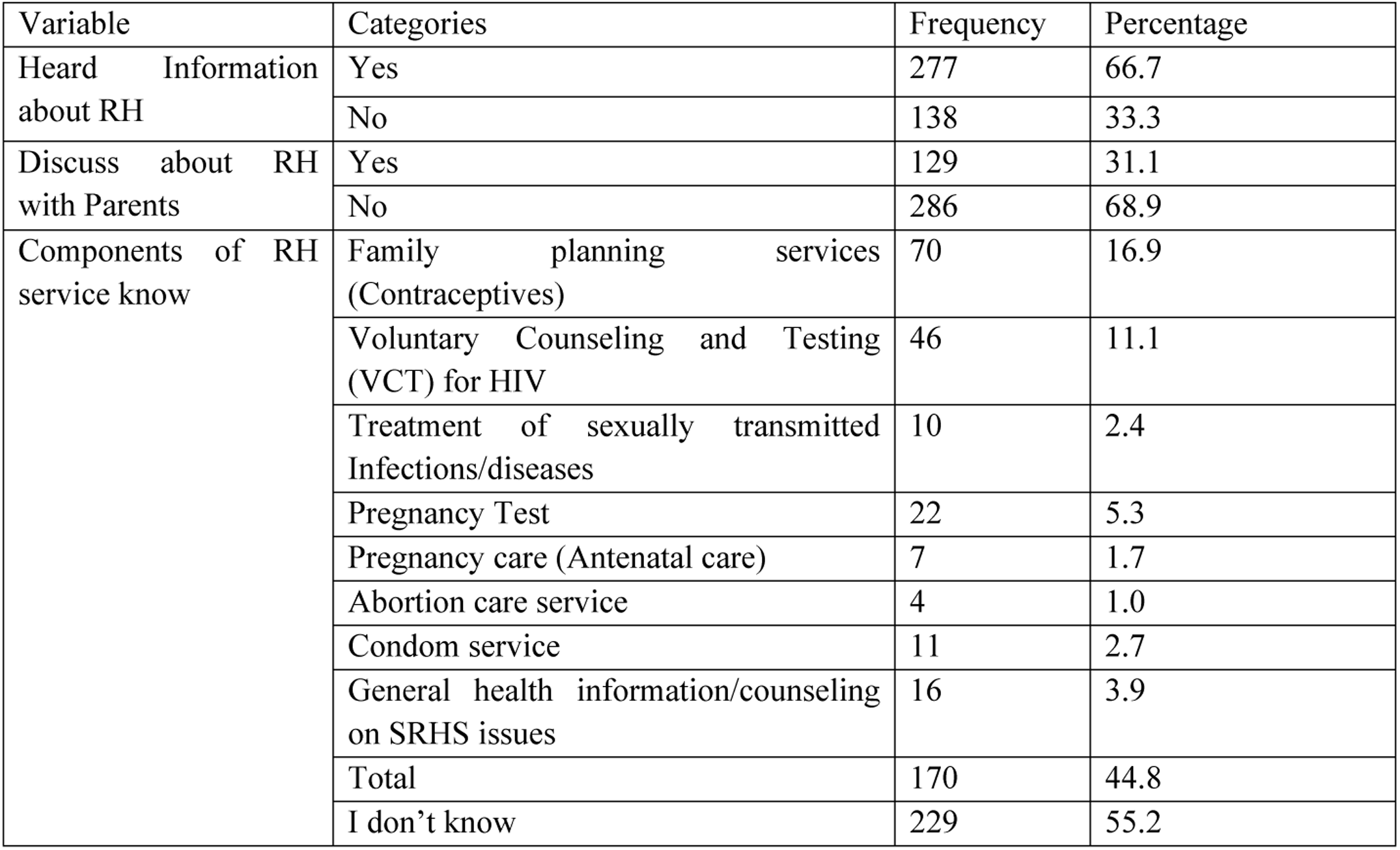
Knowledge-related characteristics of YFRHS Utilization, Yeka Sub-City, Addis Ababa, Ethiopia, 2024 (n=415)

| Variable | Categories | Frequency | Percentage |
| --- | --- | --- | --- |
| Heard Information about RH | Yes | 277 | 66.7 |
|  | No | 138 | 33.3 |
| Discuss about RH with Parents | Yes | 129 | 31.1 |
|  | No | 286 | 68.9 |
| Components of RH service know | Family planning services (Contraceptives) | 70 | 16.9 |
|  | Voluntary Counseling and Testing (VCT) for HIV | 46 | 11.1 |
|  | Treatment of sexually transmitted Infections/diseases | 10 | 2.4 |
|  | Pregnancy Test | 22 | 5.3 |
|  | Pregnancy care (Antenatal care) | 7 | 1.7 |
|  | Abortion care service | 4 | 1.0 |
|  | Condom service | 11 | 2.7 |
|  | General health information/counseling on SRHS issues | 16 | 3.9 |
|  | Total | 170 | 44.8 |
|  | I don't know | 229 | 55.2 |

As shown in the above table approximately 67 % of the participants had ever heard of youth reproductive health services and approximately 31% of the participants had a habit of discussing with their parents. Family planning services (contraceptives), Voluntary Counseling and Testing (VCT) for HIV, and Pregnancy tests were the three most well-known components of reproductive health services for youths and students, respectively.

**Figure 2:**
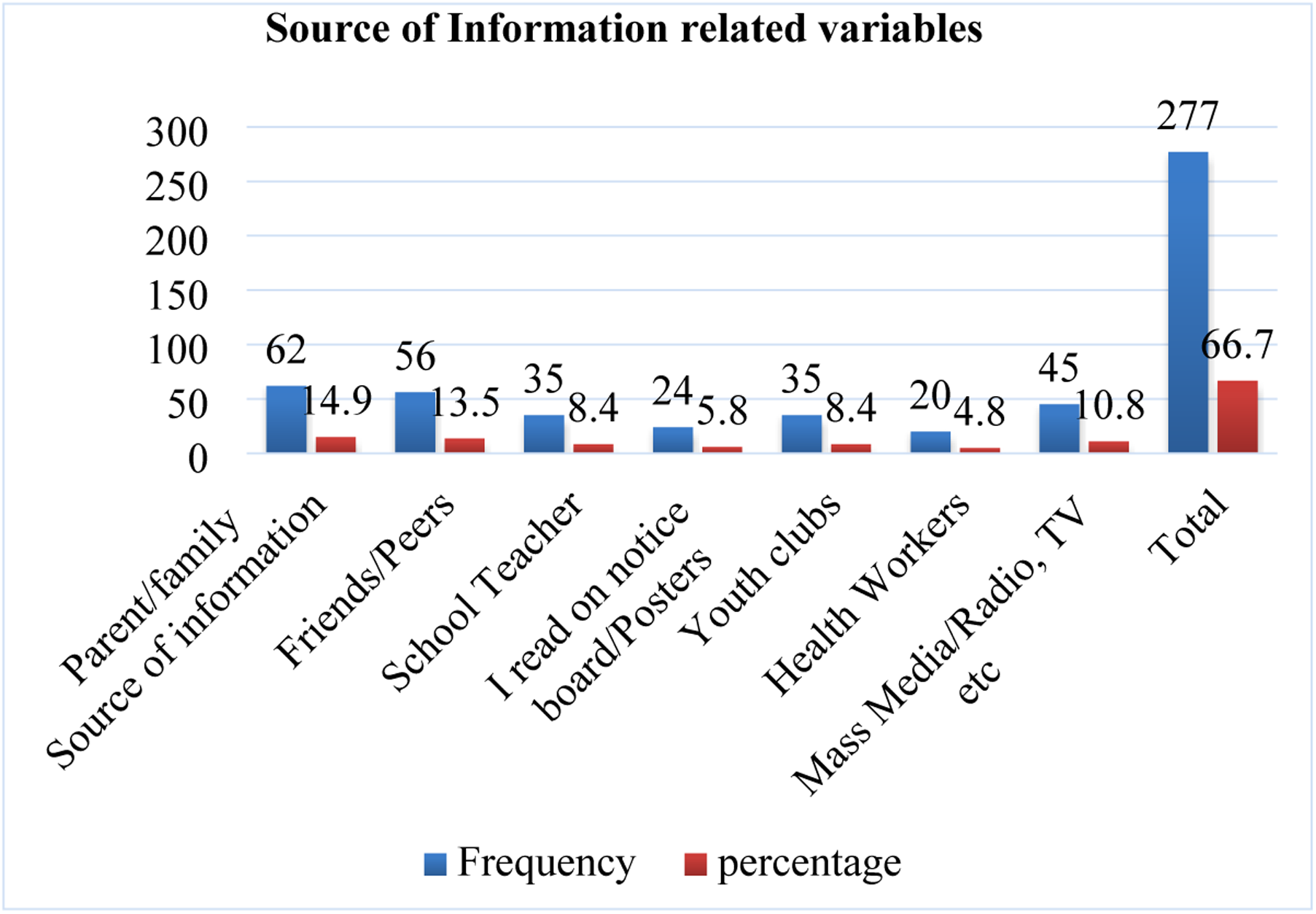
Source of information related to characteristics of YFRHS utilization, Yeka Sub-City, Addis Ababa, Ethiopia, 2024 (n=415)

As shown in the above figure parents of students, friends-/peers, mass media, and school teachers and youth clubs were the most common sources of information for students.

**Table 4:** Health-related characteristics of YFRHS Utilization, Yeka Sub-City, Addis Ababa, Ethiopia, 2024 (n=415)

| Variable | Categories | Frequency | Percentage |
| --- | --- | --- | --- |
| Availability of YFRH canter in their catchment | Yes | 173 | 41.7 |
|  | No | 242 | 58.3 |
| know the place where YFRHS found | Yes | 145 | 34.9 |
|  | No | 270 | 65.1 |
| YFRHS Improves Youths' Health? | Yes | 302 | 72.8 |
|  | No | 113 | 27.2 |
| Is Work Hour Convenient? | Yes | 128 | 74.0 |
|  | No | 45 | 26.0 |
| If no, the most convenient time for user | Earlier in the morning | 14 | 31.1 |
|  | Late in the afternoon | 12 | 26.7 |
|  | In the hours when other users not around | 19 | 42.2 |
| Preferred RH Provider | Young provider of any sex | 87 | 21.0 |
|  | Young provider of the same sex | 119 | 28.7 |
|  | Adult provider of the same sex | 82 | 19.8 |
|  | Any provider could be | 127 | 30.6 |

Regarding the type of service used by students in the last 12 months, as shown in the next table, family planning, voluntary counseling and testing (VCT) for HIV and General health information/counseling on SRHS issues were the three most common types of services. Whereas the most common reasons not to utilize YFRHSs were “Have not encountered a problem” 150 (36.1%), “Fear of being seen by parents or people whom they knew” 47 (11.3%), and “Don’t have SRHS information” 35 (8.4%) respectively.

**Table 5:**
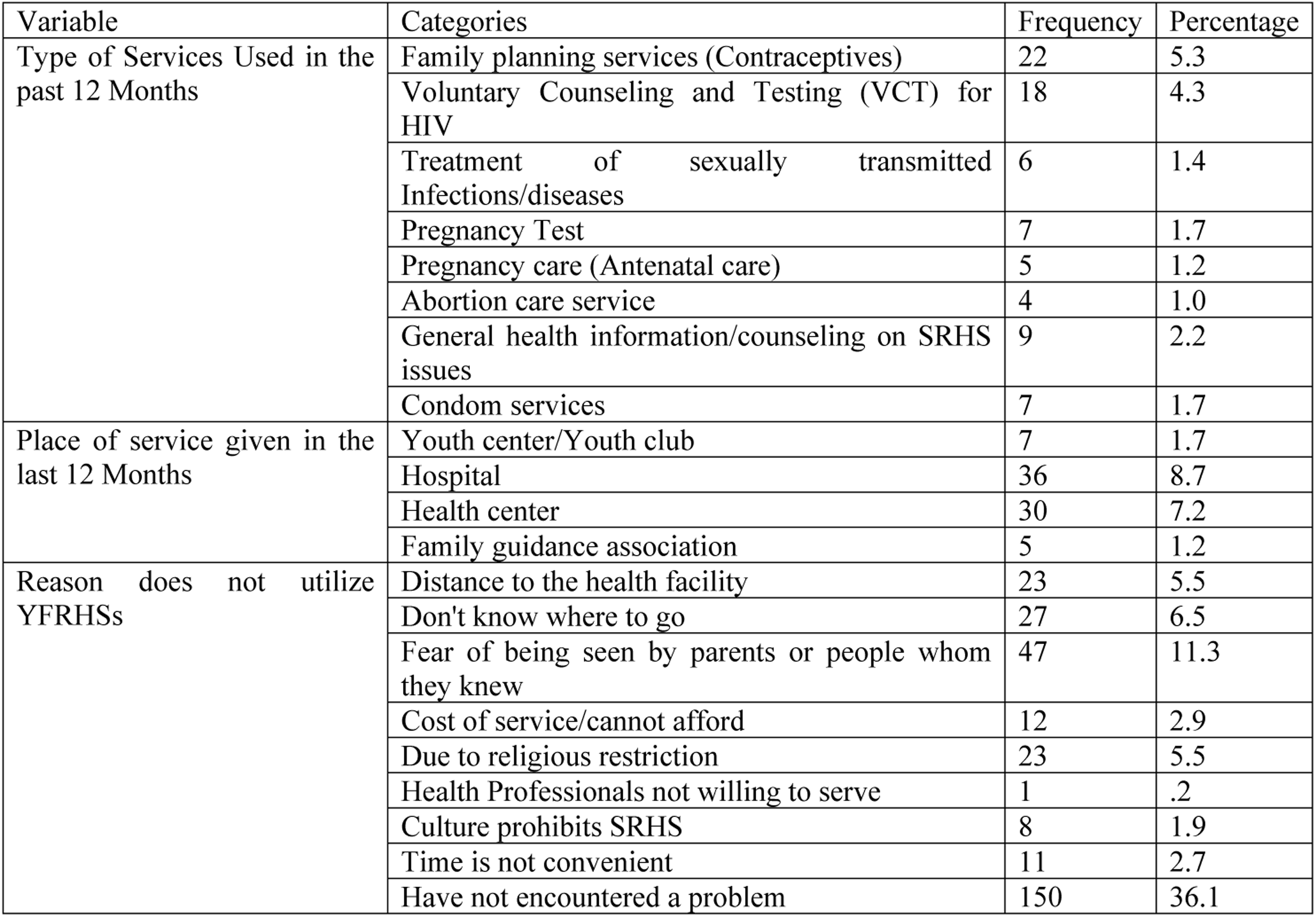
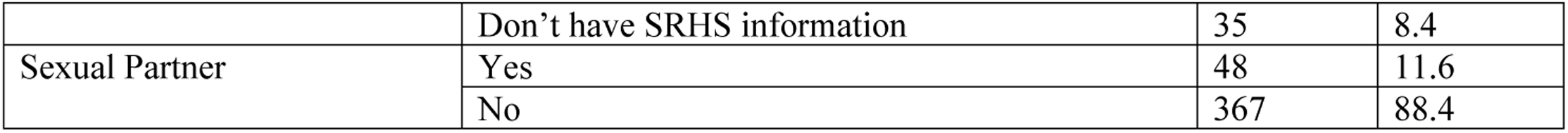
Type and Place related characteristics of YFRHS utilization, Yeka Sub-City, Addis Ababa, Ethiopia, 2024 (n=415)

| Variable | Categories | Frequency | Percentage |
| --- | --- | --- | --- |
| Type of Services Used in the past 12 Months | Family planning services (Contraceptives) | 22 | 5.3 |
|  | Voluntary Counseling and Testing (VCT) for HIV | 18 | 4.3 |
|  | Treatment of sexually transmitted Infections/diseases | 6 | 1.4 |
|  | Pregnancy Test | 7 | 1.7 |
|  | Pregnancy care (Antenatal care) | 5 | 1.2 |
|  | Abortion care service | 4 | 1.0 |
|  | General health information/counseling on SRHS issues | 9 | 2.2 |
|  | Condom services | 7 | 1.7 |
| Place of service given in the last 12 Months | Youth center/Youth club | 7 | 1.7 |
|  | Hospital | 36 | 8.7 |
|  | Health center | 30 | 7.2 |
|  | Family guidance association | 5 | 1.2 |
| Reason does not utilize YFRHSs | Distance to the health facility | 23 | 5.5 |
|  | Don't know where to go | 27 | 6.5 |
|  | Fear of being seen by parents or people whom they knew | 47 | 11.3 |
|  | Cost of service/cannot afford | 12 | 2.9 |
|  | Due to religious restriction | 23 | 5.5 |
|  | Health Professionals not willing to serve | 1 | .2 |
|  | Culture prohibits SRHS | 8 | 1.9 |
|  | Time is not convenient | 11 | 2.7 |
|  | Have not encountered a problem | 150 | 36.1 |
|  | Don't have SRHS information | 35 | 8.4 |
| Sexual Partner | Yes | 48 | 11.6 |
|  | No | 367 | 88.4 |

### School-Based Youth-Friendly Reproductive Health Service Utilization

The overall YFRHS utilization in this study was 78 (18.8 %) (95% CI: .154, .227 ).

**Figure 3:**
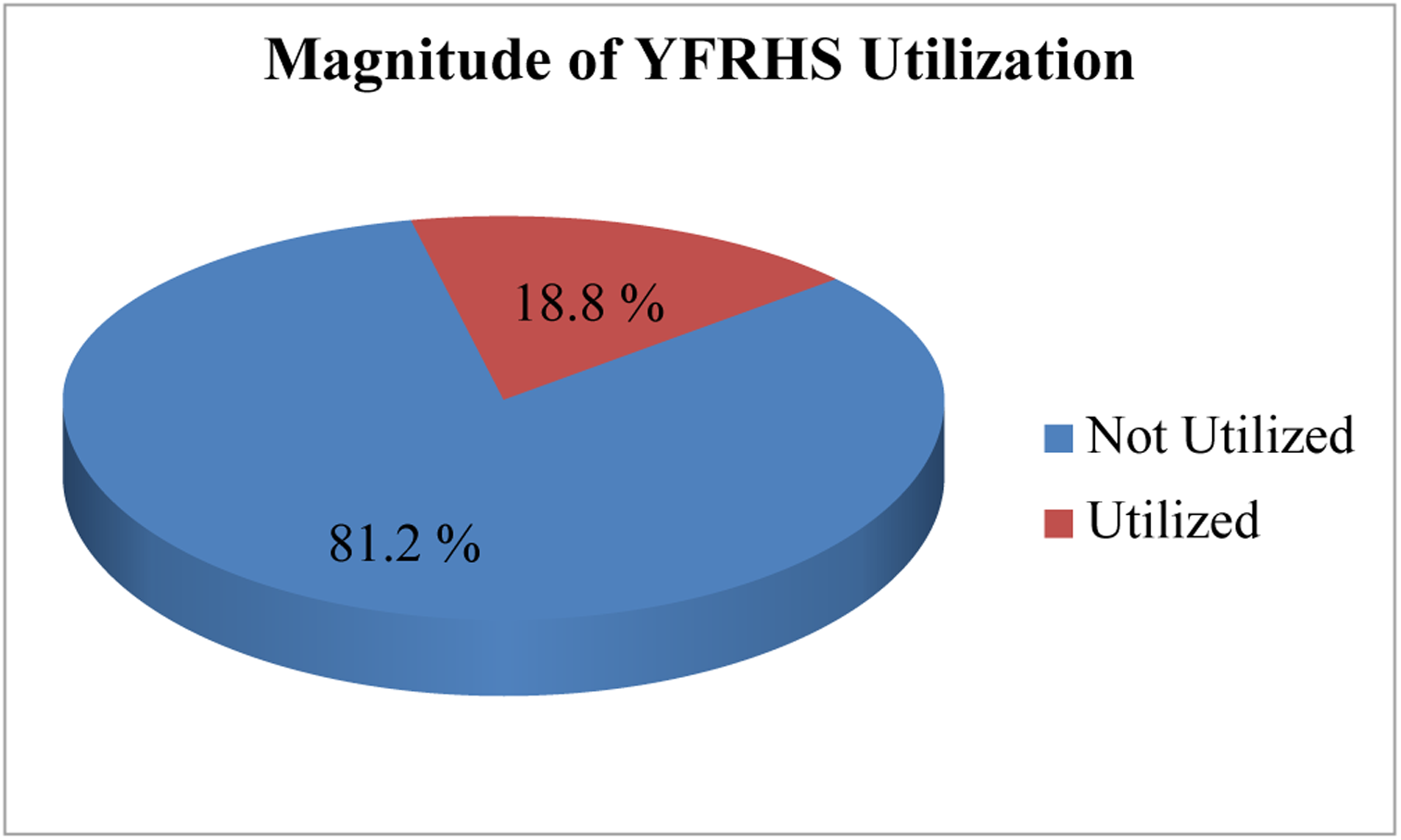
Youth-Friendly Reproductive Health Service Utilization among secondary school students of Yeka Sub-City, Addis Ababa, Ethiopia, 2024 (n = 415)

### Multivariate Analysis of Factors Associated with YFRHS Utilization

Logistic regression was performed to ascertain the effects of maternal education, discussions with parents, general information on RH, Availability of youth service centers in the living areas of the participants, knowledge of whether seeking youth centers improves youths’ health, and having sexual partners on the likelihood of utilizing RH services. The logistic regression was statistically significant. Omnibus tests of Model Coefficients less than 0. 001. The overall goodness of fit was checked using the Hosmer-Lemshow test. The predictive value of the model was assessed using the Hosmer-Lemeshow goodness-of-fit test statistics, which was found to be 0.848. Thus, the p-value for the Hosmer-Lemeshow chi-square was greater than 0.05. The values from 23.3% to 37.5% of the variation in YFRHS utilization were explained by the model and correctly classified 81.4% of cases. Discussion with parents, availability of youth service centers in the living areas of the participants, and knowledge of whether seeking youth centers improves youths’ health and having sexual partners significantly determined respondents’ YFRHS utilization.

**Table 6:** Bivariate and Multivariate Analysis of factors associated with YFRHS utilization among secondary school students of Yeka Sub-City, Addis Ababa, Ethiopia, 2024.

| Variables | Categories | YFRHS Utilization |  | COR (95% CI) | AOR (95% CI) |
| --- | --- | --- | --- | --- | --- |
|  |  | No (0) | Yes (1) |  |  |
|  |  | 337 | 78 |  |  |
| Mother's Educational Status | No formal education | 68 (20.2%) | 8 (10.3%) | 1 | 1 |
|  | Primary | 104 (30.8%) | 16 (20.5%) | 1.495 (.584, 3.823) | 2.348 (.969, 5.693) |
|  | Secondary and above | 165 (49%) | 54 (69.2%) | 2.782 (1.257, 6.157) | 2.348 (.969, 5.693) |
| Discuss about RH with Parents | No | 254 (75.4%) | 32 (41%) | 1 | 1 |
|  | Yes | 83 (24.6%) | 46 (59%) | 4.399 (2.629, 7.361) | **2.222 (1.229, 4.018)** |
| Heard Information | No | 133 (39.5%) | 5 (6.4%) | 1 | 1 |
|  | Yes | 204 (60.5%) | 73 (93.6%) | 9.519 (3.748, 24.173) | 2.551 (.886, 7.349) |
| YFRHS facility in living area (availability) | No | 228 (67.7%) | 14 (17.9%) | 1 | 1 |
|  | Yes | 109 (32.3%) | 64 (82.1%) | 9.562 (5.135, 17.806) | **5.065 (2.500, 10.260)** |
| Thought (Perception) to YFRHS improve youths' health | No | 109 (32%) | 4 (5%) | 1 | 1 |
|  | Yes | 228 (68%) | 74 (95%) | 8.844 (3.152, 24.815) | **4.595(1.504,14.039)** |
| Have sexual Partner | No | 308 (91.4%) | 59 (75.6%) | 1 |  |
|  | Yes | 29 (8.6%) | 19 (24.4%) | 3.420 (1.800, 6.500) | **3.802 (1.722, 8.395)** |

## Discussion

This study was conducted to assess the utilization of Youth-Friendly Reproductive Health Services (YFRHSs) and its associated factors among secondary school students in Yeka Sub-City, Addis Ababa, Ethiopia, in 2024. The utilization of YFRHSs in the study area was 18.8 % which was relatively lower when it was compared with Ethiopia’s pooled prevalence of youth-friendly sexual and reproductive health service utilization (13). Discussion about RH with parents, Availability of YFRHSs facilities in living areas, YFRHSs improving youths’ health (perception), and sexual partners were the determining factors.

This finding was lower than that community-based study conducted in Mandalay City, Myanmar (67%) and an institutional-based study conducted in Ghana (55.8%) and Nigeria (51%). The difference in results may be due to methodological differences in the study, socio-economic factors, infrastructure, availability, and accessibility of health facilities, and residence differences among these countries in the study area which can influence the health delivery system (14–16).

This finding was also lower compared with those similar studies conducted in Bale zone (46%), Awabel district in Amhara Region (41%), Mareka District in Dawuro zone (69.7%), Hadiya zone (38.5%), Goba town (71.4%), Mekelle (69.1%), Harar (64%) and Aleta Wondo town, Southern Ethiopia (32.8%) respectively (12, 17–22). This difference might be due to the availability and accessibility of youth-friendly RH facilities or the availability of youth centers, parenting situations, and individual/personal characteristics of the study participants.

YFRHS utilization in this study was slightly closer to a similar study conducted in the Machakal District at 21.5%, Northwest, Ethiopia, and Nekemte, Western Ethiopia, at 21.2% (23, 24). However, YFRHS utilization in this study was greater than similar study Mecha District, Northwest Ethiopia, at 18% (25) and Nepal at 9.2% (26).

The commonly utilized RH service components in this study were Family Planning/Contraceptives 22 (5.3%), voluntary counseling and testing for HIV 18 (4.3%), and General Health information/counseling on SRHS issues 9 (2.2%) respectively. The main sources of information in this study were Parent/Family 62 (14.9%), Friends/Peers 56 (13.5%), Mass Media 45 (10.8%), Youth clubs 35 (8.4%), schoolteachers 35 (8.4%), Notice board 24 (5.8%), Health workers 20 (4.8%) respectively.

The major reasons not to utilize YFRHSs for the respondents were: “Have not encountered a problem” 150 (36.1%), “Fear of being seen by parents or people when they know” 47 (11.3%), and “Don’t have SRHS information” 35 (8.4%) respectively.

In multivariable analysis, discussing RH with parents, Availability of YFRHSs facilities in living areas, Thought YFRHSs Improve youths’ health (perception), and sexual partners were positively associated with the utilization of YFRHSs.

In this study finding, respondents who had ever discussed with their parents were more than 2 times more likely to use YFRHSs than those who had not ever discussed with their parents (AOR=2.222, 95% CI: 1.229, 4.018). This might be because discussions on RH issues providing access to information about RH services. Parental discussions about sexual and reproductive issues were significantly associated with YFRHS utilization. Students who had never discussed reproductive and sexual health issues with their parents/friends were less likely to use the YFRHS than their counterparts. This study is in line with the studies conducted in Southern Ethiopia (AOR=3.17, 95% CI: 1.624, 6.206)(27), systematic reviews and meta-analyses conducted in Ethiopia (AOR=2.99, 95% CI: 1.89–4.72) (13) and Yirgalem Town, Ethiopia (AOR=4.35, 95% CI: 2.28, 8.31) (28). This may be because open discussions about SRH issues between families and their children increase awareness and avoid feeling shy and fearful while receiving SRH services. Moreover, the discussion creates more opportunities to share SRH information and experience health-related problems, and the youth would have better knowledge and awareness about SRH services and develop positive attitudes towards YFRHS which in turn might motivate them to use those services (29, 30).

In this study, the respondents who had sexual partners were almost four times more likely to use YFRHSs than their counterparts were (AOR=3.802, 95% CI: 1.722, 8.395). This finding is supported by a study conducted in Lira City West, Northern Uganda (AOR=10.0, 95% CI: 4.05-24.69)(31). A study conducted in Western Ethiopia (AOR=5.33, 95% CI: 2.53, 11.23) shows participants who had ever had ever practised sexual intercourse practice were more likely to use YFRH services compared to those who did not practice it (32). This might be because those with experience of sexual intercourse might visit health facilities either to get condoms, contraception, and HIV/AIDS counseling and testing, or to receive interventions such as abortion services when they experience negative consequences from unprotected sexual practice. This finding is in line with other similar studies conducted in Awabel District, Northwest Ethiopia (AOR=1.88, 95% CI: 1.30, 2.71) (33), Lira City West, Northern Uganda (AOR=4.59, 95% CI: 2.18-9.69)(31), and Dire Dawa City, Eastern Ethiopia (AOR = 1.95, 95% CI: 1.10, 3.44)(34), which sexually active youths were more likely to use the youth-friendly services. This might be due to sexual partners discussing SRH issues and plans for protection prior to intercourse. In addition, as a way of finding solutions to sexual and reproductive health problems, they may be likely to utilize SRH services.

This study also indicated that utilization of youth-friendly reproductive health services was high among respondents who had youth-friendly reproductive health service facilities around their living area as those who had the services in their living area were more than five times more likely to utilize youth-friendly reproductive health service as compared to those who had no youth-friendly service facilities in their living areas [AOR=5.065, 95% CI: 2.5, 10.26). This was probably due to the fact that those respondents who have the opportunity to access health facilities around their lives can be exposed to health education related to youth-friendly reproductive health services at the time of the visit, thus gaining access to learn about the types, benefits and availabilities of youth-friendly reproductive health service facilities around their lives. This study is in line with a study Ambo Town, Oromia Regional State, Ethiopia (AOR=7.7, 95% CI: 2.931-20.423) (35). Supporting this finding, a study conducted in Nigeria (36) revealed that the more a patient lives from a health facility, the less likely they are to use services. A study in Kenya also identified long distances from a facility or care source as a barrier to the use of health services (37). This is because the preferred care source is often the closest. Moreover, in the African context, the principal barriers to accessibility are transport and cost; therefore, distance is mostly reported as a single obstacle to the utilization of healthcare services (38, 39).

In this study, it was found that respondents who thought (perception) youth-friendly reproductive health services improve youths’ health more than four times utilize YFRHSs as compared to those who had no thought (perception) that youth-friendly reproductive health services didn’t improve youths’ health (AOR=4.595, 95% CI: 1.504, 14.039). This is a novel finding of the present study.

### Limitation of the Study

Due to the nature of the cross-sectional study, it may not show cause-and-effect relationships or temporal relationships. This study did not use a qualitative method to complement the quantitative results. This study was limited to institution-based studies, owing to time and resource constraints.

## Conclusion & Recommendation

In this study youth-friendly reproductive health service utilization was 18.8 % which was relatively lower when it was compared with Ethiopia’s pooled prevalence of youth-friendly sexual and reproductive health service utilization. Discussion about RH with parents, Availability of YFRHSs facilities in living areas, YFRHSs improving youths’ health (perception), and sexual partners were the determining factors. Therefore attention must be paid to the: Provide on-school or easily accessible reproductive health services, such as confidential counseling, contraceptive distribution, and STI testing and treatment. Ensuring the accessibility and availability of Reproductive Health Services. Foster an environment that de-stigmatizes discussions about reproductive health and normalizes access to related services. Provide training for teachers and staff to promote a shame-free, non-judgmental approach.

Ensure that strict confidentiality policies are in place, and that students feel secure that their personal information and health decisions will be protected. Engaging parents and caregivers in the process, provide educational resources and opportunities for open dialogue on reproductive health. Encouraging a supportive home environment. Collaborate with local healthcare providers, youth organizations, and community groups to expand the availability and accessibility of reproductive health resources for students. A comprehensive, inclusive, and student-centered approach should be adopted to promote the utilization of reproductive health services and empower young people to make informed decisions regarding their sexual and reproductive health.

## Data Availability

N/A

## Abbreviations

ANC: Antenatal Care
RH: Reproductive Health
STIS: Sexually Transmitted Infections
UNFPA: United Nations Fund for Population Activities
YFRHS: Youth-Friendly Reproductive Health Service
WHO: World Health Organization.

## Acknowledgment

We would like to express our sincere gratitude to the data collectors, supervisors, and all students who participated in this study.

## Author’s contribution

All authors contributed towards and agreed to be accountable for all aspects of this work. All authors contributed equally contributed to data analysis, drafting, and revision of the manuscript. The final version of the manuscript was approved by all authors.

## Availability of data and materials

The datasets used and/or analyzed during the current study are available from the corresponding author upon reasonable request.

## Ethics approval and consent to participate

Ethical clearance was obtained from the Addis Ababa Public Health and Emergency Management Directorate and ethics referral No_ A/A/19725/227. The study participants provided written informed assent and consent to participate after receiving information about the purpose of the study, risks and benefits, and their rights. Assurance of privacy during the interview and confidentiality of information were provided.

## Consent for publication

“Not applicable”.

